# Vascular flow simulations using SimVascular and OpenFOAM

**DOI:** 10.1101/2021.09.11.21263191

**Authors:** Swetha Yogeswaran, Fei Liu

## Abstract

Applications of computational fluid dynamics (CFD) techniques to aid in the diagnosis and treatment of cardiovascular disease have entered the research domain in recent years, due to their ability to provide valuable patient-specific information without risks associated with highly invasive procedures. SimVascular [1] [2] is an open-source software which allows streamlined processing and CFD blood flow analysis of medical imaging data. OpenFOAM [3] is a proven open-source software which allows for versatile modeling of various fluid dynamics phenomena. In this study, both SimVascular and OpenFOAM simulations are set up with identical computational mesh, similar numerical schemes, boundary conditions, and material properties, to model blood flow in the coronary artery of a 10 year old patient with Coarctation of the Aorta (CoA) who underwent end-to-side anastomosis. Difference in the flow fields such as flow rate, pressure, vorticity, and wall shear stress between SimVascular and OpenFOAM are analyzed. Similar results are obtained in both simulations up to a certain model time, before the results become drastically different. Both the similarities and differences are documented and discussed.

## 1 Introduction

CFD is a well-established technique in modeling heat, mass, and momentum transfer, which is especially applicable in many areas of engineering, including aerospace [13], automotive [14], and biomedical [15]. Specifically, CFD simulations can play a significant role in the development and testing of products, as it allows engineers to understand fluid interaction with products without the expensive and time consuming process of physically building and testing. More recently, CFD applications in cardiology have been explored extensively, as they can not only obviate the need for risky invasive measurement techniques, but also provide provide measurements of particular hemodynamic parameters that can not otherwise be measured directly, such as wall shear stress (WSS). This is also a step towards more personalized treatments, since patient-specific models can give insight as to what treatment plans may be more effective for the specific case, as opposed to just the general population [15]. To run these simulations, SimVascular, an open-source software, provides a very streamlined process to run blood flow simulations, starting from medical images. Because of its specialized nature, it allows for particular cardiology-specific customizations, such as RCR boundary conditions, to be applied in an easy, straightforward way. On the other hand, Open-FOAM, a well-established CFD software, allows for versatile modeling of various fluid dynamics phenomena, and has a wide variety of customizations for many different applications. Although there are a variety of CFD software that may each have different benefits, it is important that final results are very similar when a particular case is modelled, as the planning of treatment strategies can only be aided by CFD simulations if the simulations are consistent and reliable. So, in this work, simulations are run on both SimVascular and OpenFOAM in order to better understand the effects of the CFD software on subsequent results.

## 2 Data

This study uses data from the Cardiovascular and Pulmonary Model Repository from the Open Source Medical Software Corporation (OSMSC). This specific case was contributed by LaDisa et al [4]. This case shows the aorta and few surrounding vessels of a 10 year old female with Aortic Coarctation, a narrowing of the aorta, after the surgical end-to-side anastomosis procedure, where vessels were placed to connect the aorta before and after the coarctation in order to allow sufficient blood flow to the area of the aorta past the narrowing [4].

## 3 Model Setup

Where it is possible, SimVascular and Open-FOAM are set up identically. For example, the geometry and mesh of the vessel, wall boundary condition, and the flow rate at the inlet are identical. There are also inherent differences, such as numerical algorithms, pressure boundary conditions, and material model. We discuss the similarities and differences in the setup in the following subsections.

### 3.1 Geometry

### 3.2 Meshing

The 3D geometry (Figure 1) of the aortal model is taken directly from LaDisa et al [4]. This geometry is imported into SimVascular where it is meshed with a global max edge size of 0.1 cm, resulting in a mesh with 118950 nodes and 669813 cells as shown in Figure 2. The mesh from SimVascular in .vtu format is then converted to TetGen .ele and .node files using the mesh conversion function from the meshio package [5]. These TetGen files are then converted to mesh formats compatible with OpenFOAM using the tetgenToFoam function from OpenFOAM. After bringing the mesh to OpenFOAM, the mesh is scaled by a facter of 0.01, since SimVascular simulations are run in cgs units while OpenFOAM simulations are run in SI units. Finally, the different faces of the mesh are extracted using the autopatch option on OpenFOAM. After this process, identical meshes are ready for simulation on both SimVascular and OpenFOAM.

**Figure 1:**
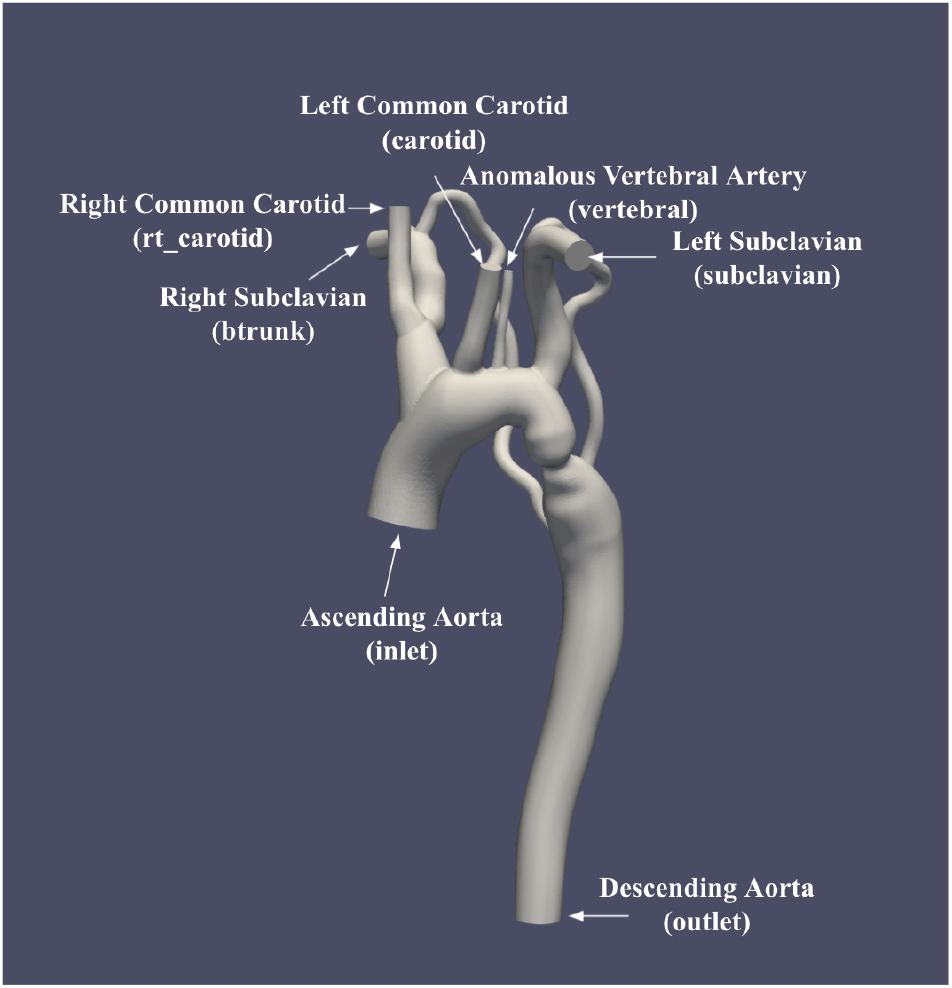
Labeled arteries and associated faces. Cross-sectional areas are provided in Table 2

**Figure 2:**
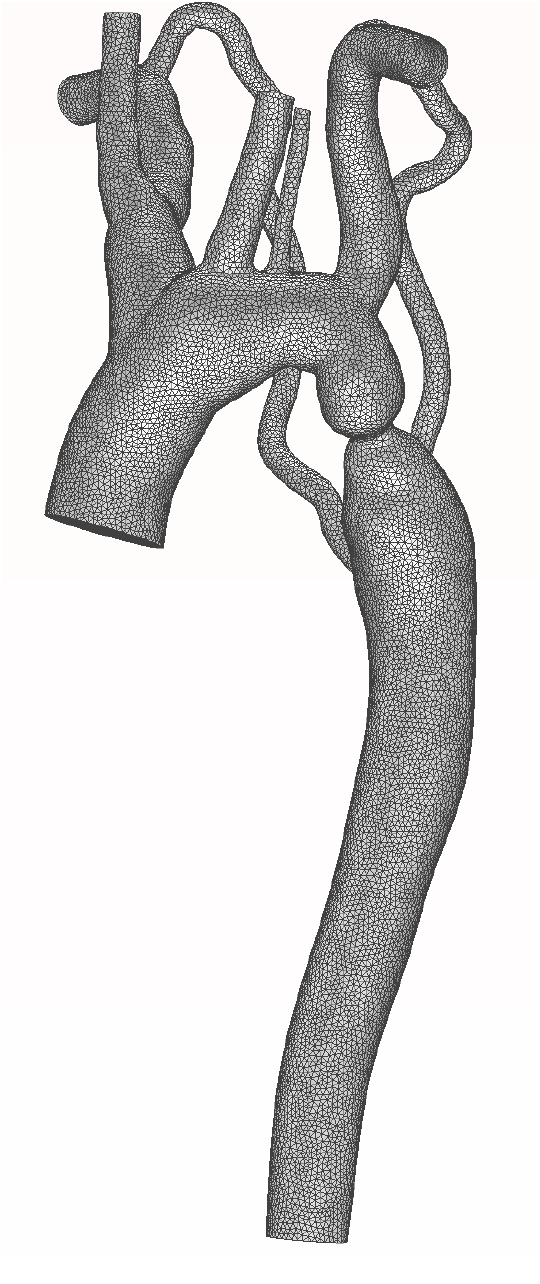
Mesh of the artery

### 3.3 Wall and Fluid Parameters

The wall is modelled as a rigid surface in both software and no-slip boundary conditions are applied on the vessel wall.

In SimVascular, fluid density and viscosity are set as default, 1.06*g* · *cm*^*−*3^ and 0.04*g* · *cm*^−1^ · *s*^−1^ respectively.

The fluid properties in OpenFOAM are based on the non-Newtonian Bird-Carreau model of blood [18] [19], where the kinetic viscosity *v* of blood varies as a function of rate of shear stress 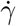 in the fluid shown in Equation 1. The exact numbers used are: *ν*_0_ = 3.3*e* − 6*m*^2^*/s, ν*_*∞*_ = 1.32*e* − 05*m*^2^*/s, K* = 0.6046*s, n* = 0.3742, *a* = 2.

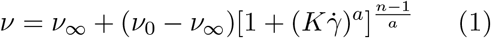

### 3.4 Boundary Conditions

#### 3.4.1 Inlet Condition

The inlet condition for one period of blood flow is read and approximated from the graph of inlet flow rate provided by the data adopted from LaDisa et al [4]. A spline smoothed flow rate interpolated from the approximated data points is applied for three periods in both SimVascular and OpenFOAM as shown in Figure 3, so the inlet flow rate boundary conditions are identical in both simulations. The pressure at inlet in both SimVascular and OpenFOAM are solved from the Navier-Stokes equations.

**Figure 3:**
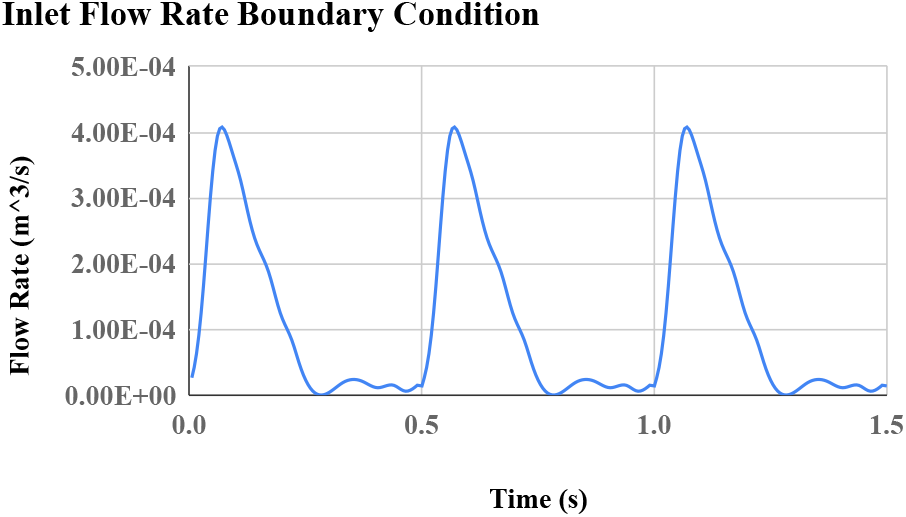
Graph of inlet boundary condition: Inlet volumetric flow rate over time

#### 3.4.2 Outlet Condition

The outlet boundary conditions are set up to be as close as possible between SimVascular and OpenFOAM.

On SimVascular, a resistance-capacitance-resistance (RCR) model is employed on all outlets, as given by LaDisa et al [4] and listed in Table 3. The RCR model allows simulating opposing pressure at the outlet boundary due to down-stream resistance from wall shear stress. The capacitance term simulates the effect of artery wall dilation and contraction modulating blood flow downstream. The RCR-associated opposing pressures are time dependent. The average pressure on each outlet for each time step is recorded from SimVascular. These resulting pressures are then spline smoothed and set as outlet pressure boundary conditions on OpenFOAM. An important detail here is the scaling of pressure when the pressure boundary is applied in Open-FOAM. OpenFOAM works with kinematic pressure scaled by fluid density 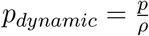. So, the pressure values from SimVascular are scaled by fluid density before applied at the outlet boundaries in OpenFOAM. In addition, because of the unit system difference between SimVascular and OpenFOAM, the pressure values from SimVascular measured in cgs are converted to SI before applied at the outlet boundaries in OpenFOAM.

**Table 1:** Total Dimensions

| Volume ($m^3$) | Surface Area ($m^2$) |
| --- | --- |
| $4.337 * 10^{-5}$ | $1.556 * 10^{-2}$ |

**Table 2:** Cross Sectional Areas of Inlets and outlets.

| Face | Cross Sectional Area ($m^2$) |
| --- | --- |
| btrunk | $4.37 * 10^{-5}$ |
| carotid | $2.14 * 10^{-5}$ |
| inlet | $2.51 * 10^{-4}$ |
| outlet | $1.07 * 10^{-4}$ |
| rtcarotid | $2.12 * 10^{-5}$ |
| subclavian | $3.48 * 10^{-5}$ |
| vertebral | $3.90 * 10^{-6}$ |

**Table 3:** RCR values employed in SimVascular in cgs units

| Face | $R_p$ | $C$ | $R_d$ |
| --- | --- | --- | --- |
| btrunk | 1102 | 0.0001024 | 18934 |
| carotid | 1396 | 0.0000932 | 20085 |
| outlet | 98 | 0.0012393 | 1088 |
| rtcarotid | 1322 | 0.0000932 | 19017 |
| subclavian | 1155 | 0.0001320 | 12847 |
| vertebral | 9681 | 0.0000166 | 111331 |

### 3.5 Solver

The governing equations of the incompressible viscous fluids can be written as

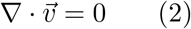

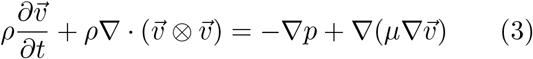

Note that because density is constant in incompressible fluids, the momentum equation is often written in terms of kinematic pressure 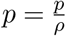 and kinematic viscosity 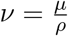.

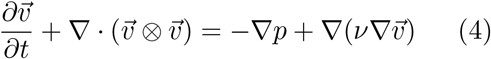

Both SimVascular and OpenFOAM solve the same set of equations. However their approaches to the solution of the Navier Stokes equations are different. SimVascular uses a finite element method while OpenFOAM uses a finite volume method. Both approaches are well equipped to work with adaptive mesh with variable resolution. This is useful where the characteristics of the blood flow is important near the wall. Past work [16], [17] have shown properties such as wall shear stress can play an important role in plaque development. Thus the ability to solve Navier Stokes equations with refined mesh elements near the vessel wall is important. Both the finite element approach in SimVascular and finite volume approach in OpenFOAM can produce detailed solutions near the vessel for further investigation. Turbulence calculation is turned off in both SimVascular and OpenFOAM.

In order to compare the predictive ability of SimVascular and OpenFOAM in simulating blood flow patterns, we have set up a case between SimVascular and OpenFOAM where the conditions of the simulation are set up in identical ways where it is possible.

The following sections discuss similarities and differences between the two different simulations.

#### 3.5.1 SimVascular

The SimVascular solver solves the same incompressible viscous fluid equations on geometry following mesh using finite element algorithm [1].

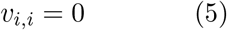

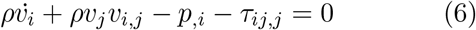

where *ρ* is blood density, *v*_*i*_ is the *i*-th component of the fluid velocity and 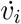 its time is derivative, *p* is the pressure, and *τ*_*ij*_ is the viscous portion of the stress tensor. The flow solver inside of SimVascular evolved from the academic finite element code PHASTA (Parallel, Hierarchical, Adaptive, Stabilized, Transient Analysis) for solving the Navier-Stokes equations in an arbitrary domain with the streamline-upwind/Petrov-Galerkin (SUPG) and pressure-stabilizing/Petrov Galerkin (PSPG) methods [12].

The SUPG/PSPG formulation is defined on the finite-dimensional trial solution and weight function spaces 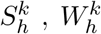, and 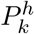 The domain is denoted by Ω ∈ ℝ^3^, and its boundary by Γ = Γ_*D*_ ∪ Γ_*N*_. Dirichlet boundary conditions are applied on Γ_*D*_, and Neumann, or flux type boundary conditions are applied on Γ_*N*_. Ω is discretized by *n*_*el*_ linear elements, Ω_*e*_.

#### 3.5.2 OpenFOAM

The OpenFOAM PISO solver uses a finite volume mesh based algorithm to solve the transient incompressible viscous fluid flow [9] [10] [11].

The pressure-velocity coupled equation is solved by decoupling the pressure and momentum fields through predictor-corrector steps. During the momentum predictor step, *H* matrix is solved from the momentum equation which has been matrixized on the finite volume mesh.

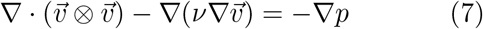

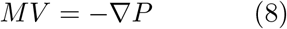

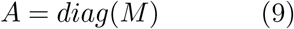

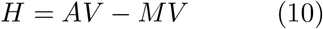

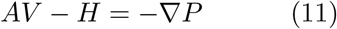

Now we can start the iterative process solving for pressure *P* and velocity *V*. Start with the momentum equation,

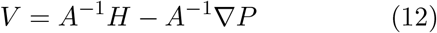

Substituting the *V* equation into the continuity equation leads to a Poisson equation of pressure that can be solved by under relaxation method.

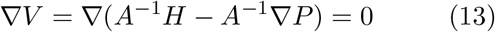

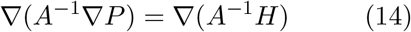

First, in the predictor step, the momentum equation is solved using initial pressure *P* and velocity boundary condition to find velocity *V*

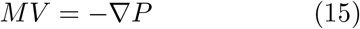

The velocity field *V* is then used in the Poisson equation to solve pressure *P*

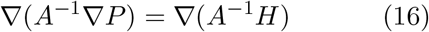

The pressure field is then used in equation (12) to correct *V* at the boundary. This is the corrector step. This process is iterated until a solution of *V* and *P* converges for the computational domain.

## 4 Results

In the following sections, we compare SimVascular and OpenFOAM results for the first 0.25 seconds. SimVascular and OpenFOAM show similar results for the first 0.25 seconds. We also discuss the differences between SimVascular and OpenFOAM after 0.25 seconds in Section 5.

### 4.1 Pressure

Due to the use of RCR parameterization (Table 3) in SimVascular, its pressure changes dynamically at the outlets. To bring the two simulations in line, the pressure values at the outlets in SimVascular are exported, converted, and applied in OpenFOAM as discussed in Section 3.4.2.

With identical outlet pressure boundary conditions and inflow rate boundary conditions, the simulated inlet pressure patterns agree very well between SimVascular and OpenFOAM (Figure 4)

**Figure 4:**
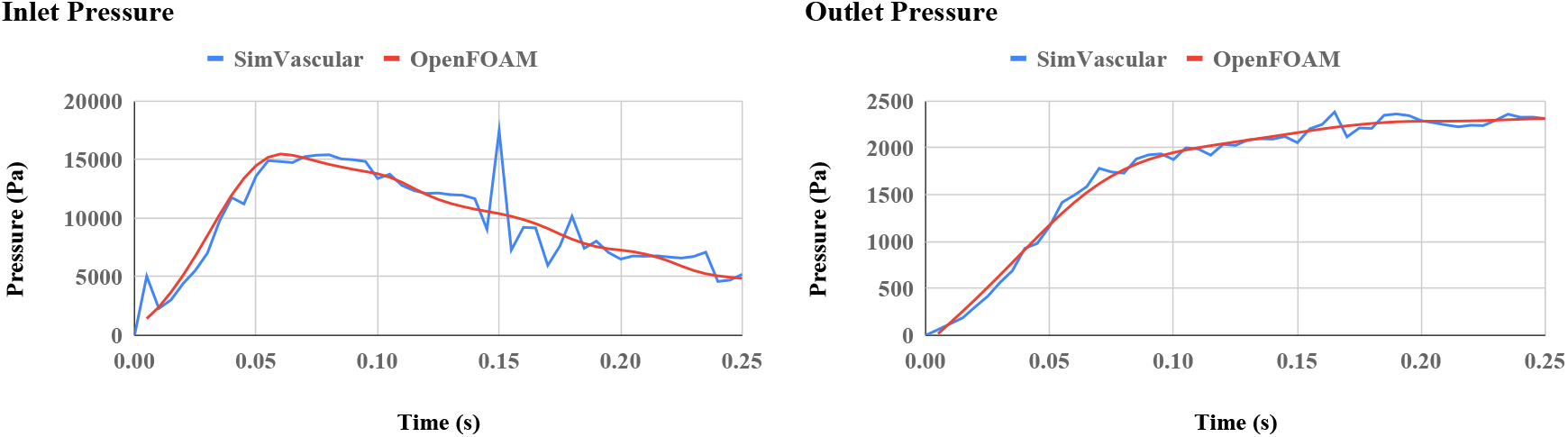
On the left: Simulated inlet pressure in SimVascular and OpenFOAM. The pressure pattern over time agrees well. On the right: Pressure values at the outlet in SimVascular and OpenFOAM. The pressure boundary condition at the outlet in OpenFOAM is prescribed from the simulated result from SimVascular, so the values match as expected.

### 4.2 Flow Rate

The total inflow and outflow rates in SimVascular and OpenFOAM (Figure 5) both show perfect agreement because both SimVascular and OpenFOAM use incompressible fluid solvers and have identical inlet flow boundary conditions. We expect this perfect agreement from both models.

**Figure 5:**
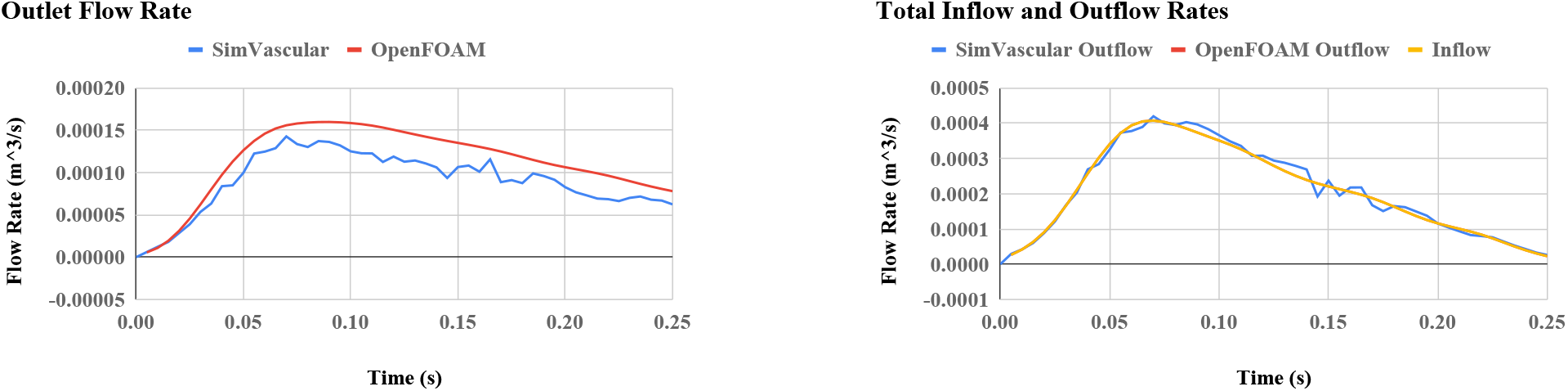
On the left: Flow rate at the outlet computed by SimVascular and OpenFOAM. On the right: Total flow rate at the outlets computed by SimVascular and OpenFOAM. The total outflow rate is the sum of outflow rates at all the outlets. The total outflow rate is compared with the flow rate at the inlet. The OpenFOAM outlet flow rate is identical to the inlet flow rate and the two curves are exactly superimposed.

The computed flow rates at the outlet show reasonable agreement. All outlets have similar agreements for the first 0.25 seconds, although some tend to have slightly higher flows in Open-FOAM while others have slightly lower flows. The flow rate at different outlets as a function of time simulated by both SimVascular and Open-FOAM are shown in Figure 6.

**Figure 6:**
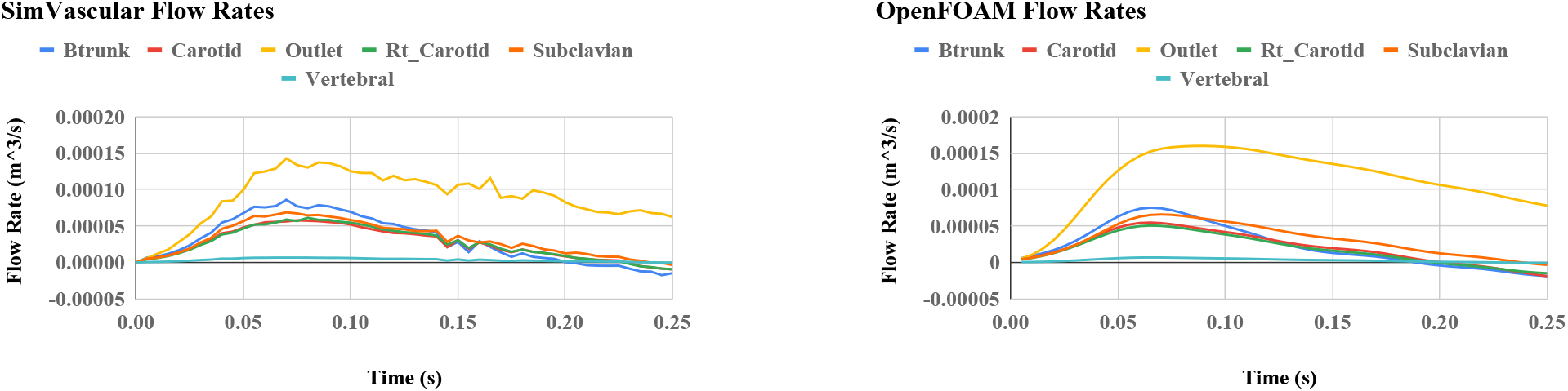
On the left: SimVascular simulated flow rates the outlets. On the right: OpenFOAM simulated flow rates at the outlets.

### 4.3 Velocity

Similar velocity magnitudes is observed in both SimVascular and OpenFOAM. Note that we have to apply a speed limiter at 7 m/s in Open-FOAM to keep the OpenFOAM solver stable. Without the speed limiter, the maximum speed modelled by OpenFOAM increases to unreasonable values. Constrained by the Courant condition, this increase in speed causes the dynamic time step to decrease to less than 10^*−*20^ seconds, which eventually halts the calculation. Therefore, we suspect the velocities modelled by OpenFOAM at certain mesh cells is higher and there may be numerical issues with the specific mesh used that causes numerical instability. The speed limiter is able to dampen the flow artificially and allows the simulation to continue. Figure 7 compares the velocity at 0.07 seconds between SimVascular and OpenFOAM visualized through streamlines within the mesh wall in Paraview [6]. It can be seen the velocity values and flow pattern modelled are in good agreement between SimVascular and OpenFOAM. Overall, the velocity in OpenFOAM is lower than what’s modelled in SimVascular, further supporting our argument for the application of the speed limiter.

**Figure 7:**
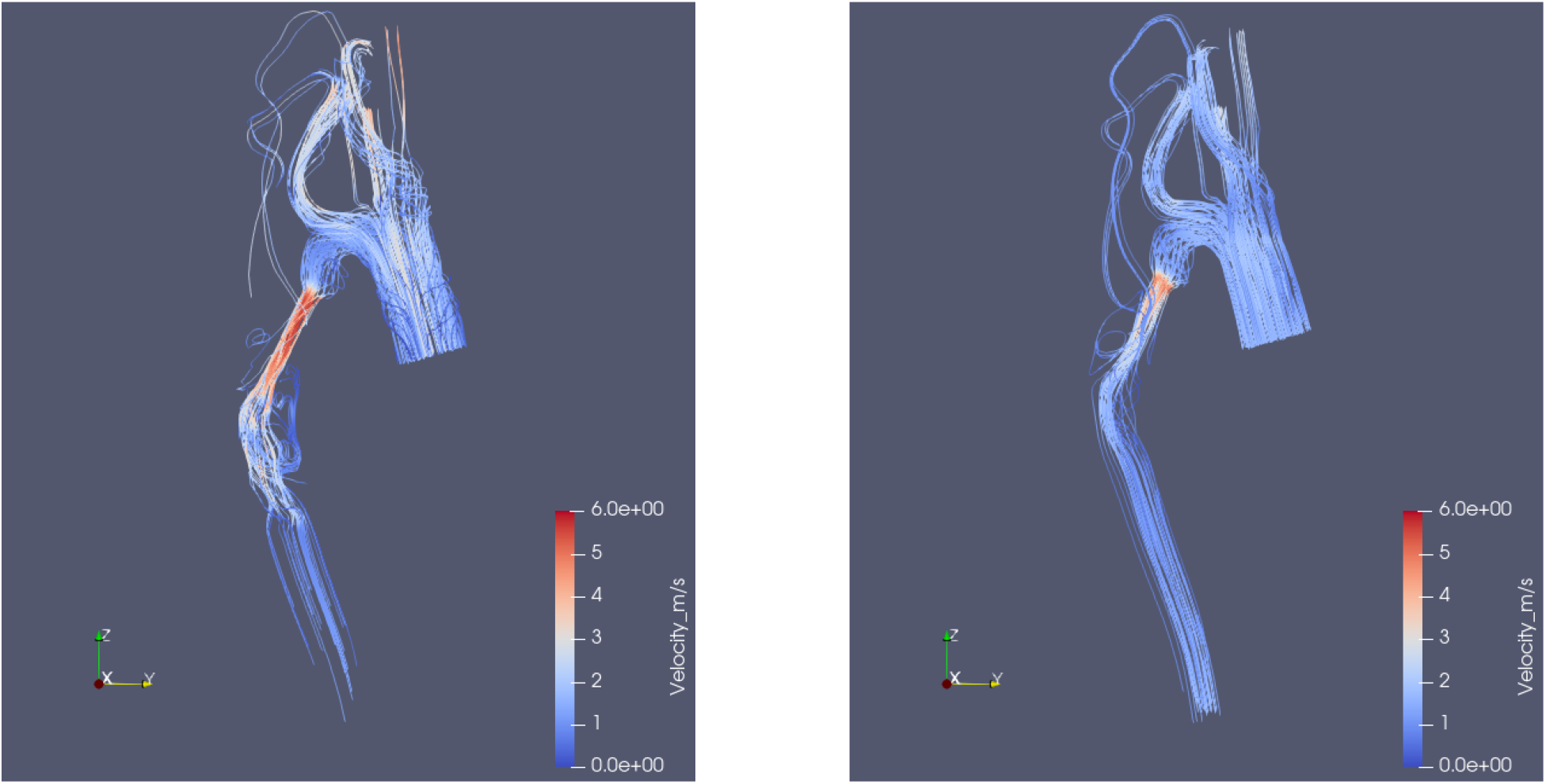
On the left: Velocity results from SimVascular at 0.07 seconds. On the right: Velocity results from OpenFOAM at 0.07 seconds. 0.07 seconds is chosen for the time to display results because it is when inflow rate is at the maximum. Note both color scales are identical, and all velocities are shown in m/s.

### 4.4 Wall Shear Stress

Although wall shear stress *τ* = *μ*∇*v* has similar qualitative patterns between SimVascular and OpenFOAM, they have very different values as seen in Figure 8. Note that the native WSS result from SimVascular is measured in dyne/cm^2^ while the OpenFOAM result is in m^2^/s^2^. After necessary conversions, both WSS results are shown in Pascals. The WSS in OpenFOAM is about one order of magnitude higher than that calculated from SimVascular. This difference is most likely caused by the non-Newtonian model of shear stress rate calculation in OpenFOAM explained in Section 3.3.

**Figure 8:**
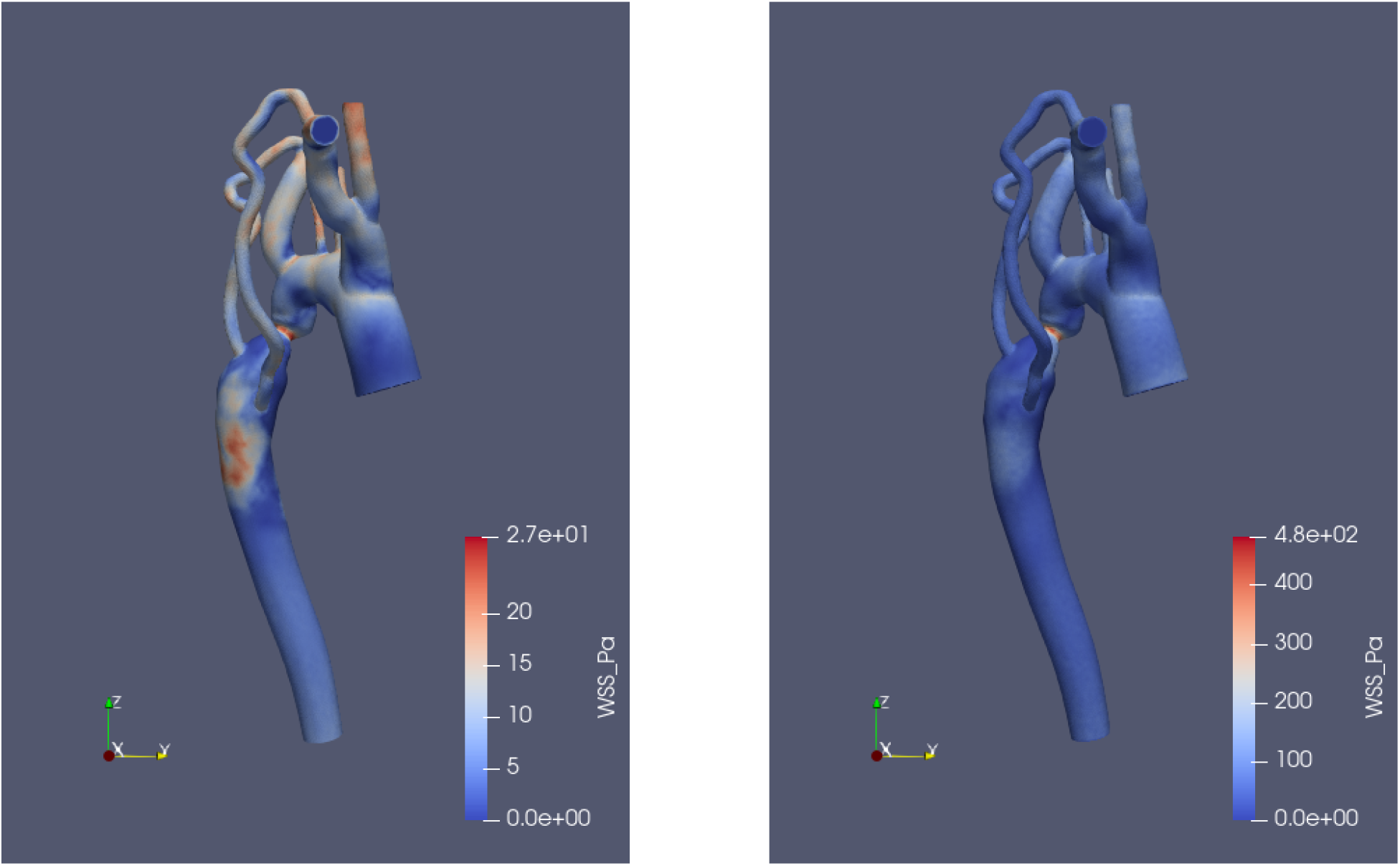
On the left: SimVascular WSS results at 0.07 seconds. On the right: OpenFOAM WSS results at 0.07 seconds. Similar qualitative patterns are observed with the WSS, but values are very different. Note the color scales are different due to drastically different values.

### 4.5 Vorticity

Similar qualitative results for vorticity are seen in SimVascular and OpenFOAM, but values differ slightly as shown in Figure 9. Mean vorticities for each timestep agree very well between SimVascular and OpenFOAM.

**Figure 9:**
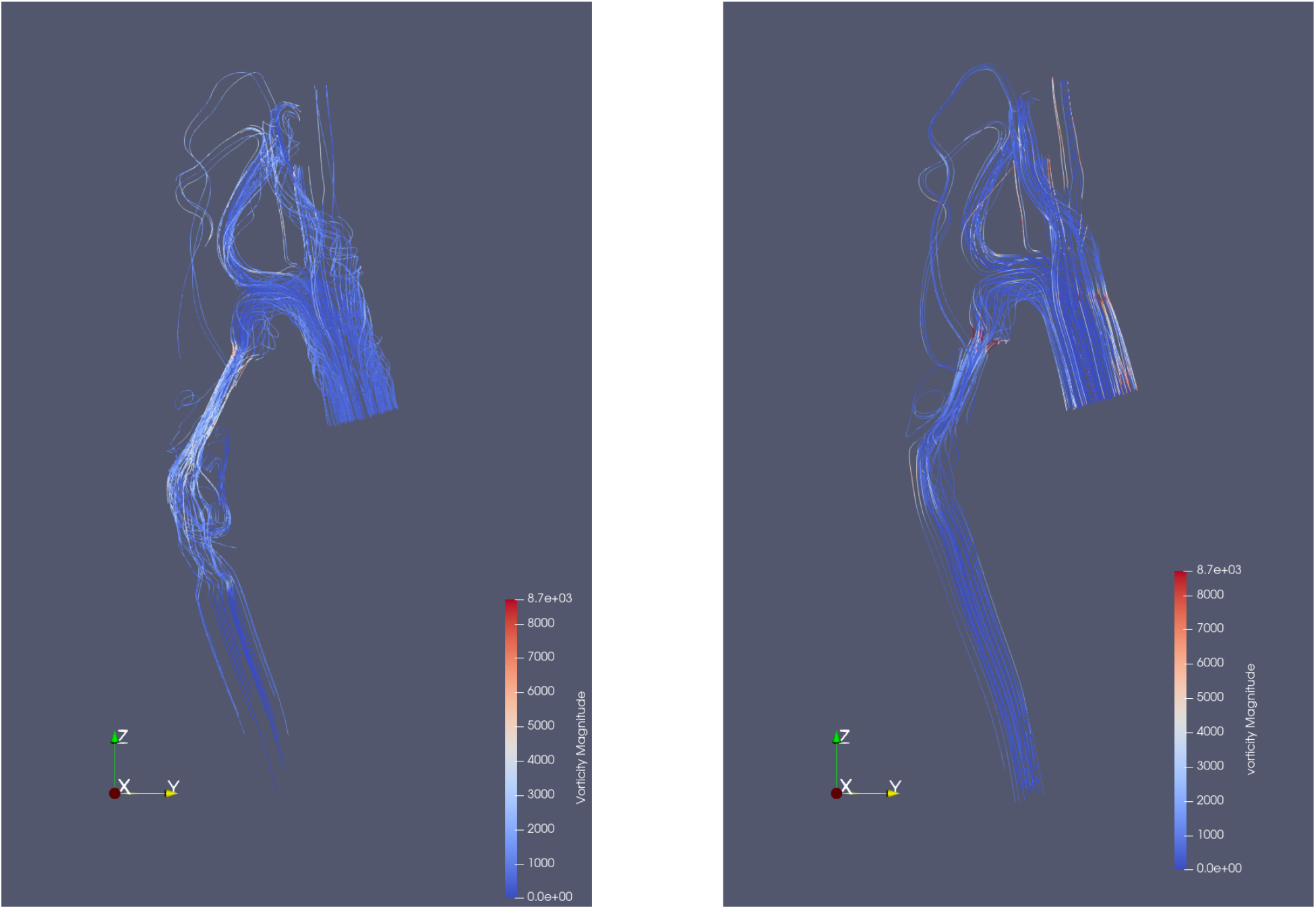
On the left: Vorticity results from SimVascular at 0.07 seconds. On the right: Vorticity results from OpenFOAM at 0.07 seconds. Similar qualitative results are shown, but values differ slightly. Color scales of both images are the same from 0 to 8700 1/s.

## 5 Discussion and Future Work

The aorta and few surrounding vessels of a patient is used to simulate blood flow patterns in both SimVascular and OpenFOAM. Other than the inherent differences in the fluid solvers, one being finite element based and one being finite volume based, geometry, meshing, and solver parameters are chosen in SimVascular and Open-FOAM to be as identical as possible. To model the effect of downstream RCR parameterization on pressure, the pressure values from SimVascular’s simulation are used directly as boundary conditions in the OpenFOAM setup. Both fluid solvers use identical inlet blood flow rates for three periods. A no-slip smooth wall boundary condition in the interior of the vessel is applied in both solvers. The fluid properties are different: SimVascular uses Newtonian fluid while the OpenFOAM fluid is non-Newtonian and simulates varying viscosity as a function of shear rate in the fluid.

The following flow fields are examined, pressure (Section 4.1), flow rate (Section 4.2), velocity (Section 4.3), WSS (Section 4.4), and vorticity (Section 4.5). Apart from WSS, both solvers show remarkably good agreement on the results simulated up to 0.25 seconds. The difference in WSS can be attributed to the difference in fluid properties used, which is discussed in Section 3.3.

Other than the impressive similarities, we discover a few noteworthy discrepancies in the SimVascular and OpenFOAM simulated results after 0.25 seconds:

1. Although the pressure values at the outlets are specified as boundary conditions, some outlets in SimVascular flip this specified pressure during simulation. This is completely unexpected, given how boundary conditions are normally applied in such solvers. The earliest flip or reversal is seen in the Carotid face at around 0.25 seconds (Figure 11), which is when the inlet flow rate decreases to essentially 0 (Figure 3). The pressure result is obtained by running post-processing utilities on the simulated result in OpenFOAM and the values are not artificial as evidenced by the velocity glyph (Figure 12). Because of this odd result, we chose to compare only the first 0.25 seconds in Section 4. Further investigation is needed to understand the cause of this reversal in pressure in OpenFOAM.

**Figure 10:**
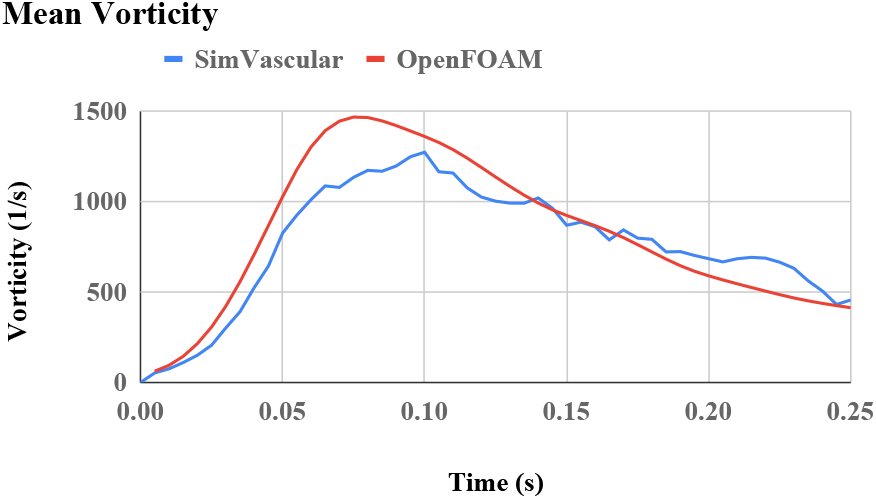
Mean vorticity calculated by both SimVascular and OpenFOAM are shown over time. The values are reasonably similar.

**Figure 11:**
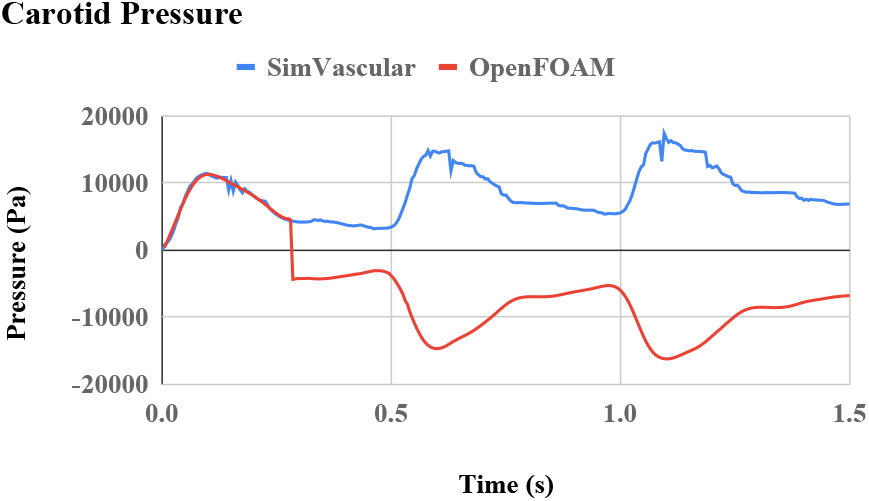
Carotid pressures over time in SimVascular and OpenFOAM are shown. Note that the pressure in SimVascular is inputted as the pressure boundary condition in OpenFOAM. At around 0.25 seconds, the OpenFOAM pressure is essentially a mirror image of the prescribed boundary conditions.

**Figure 12:**
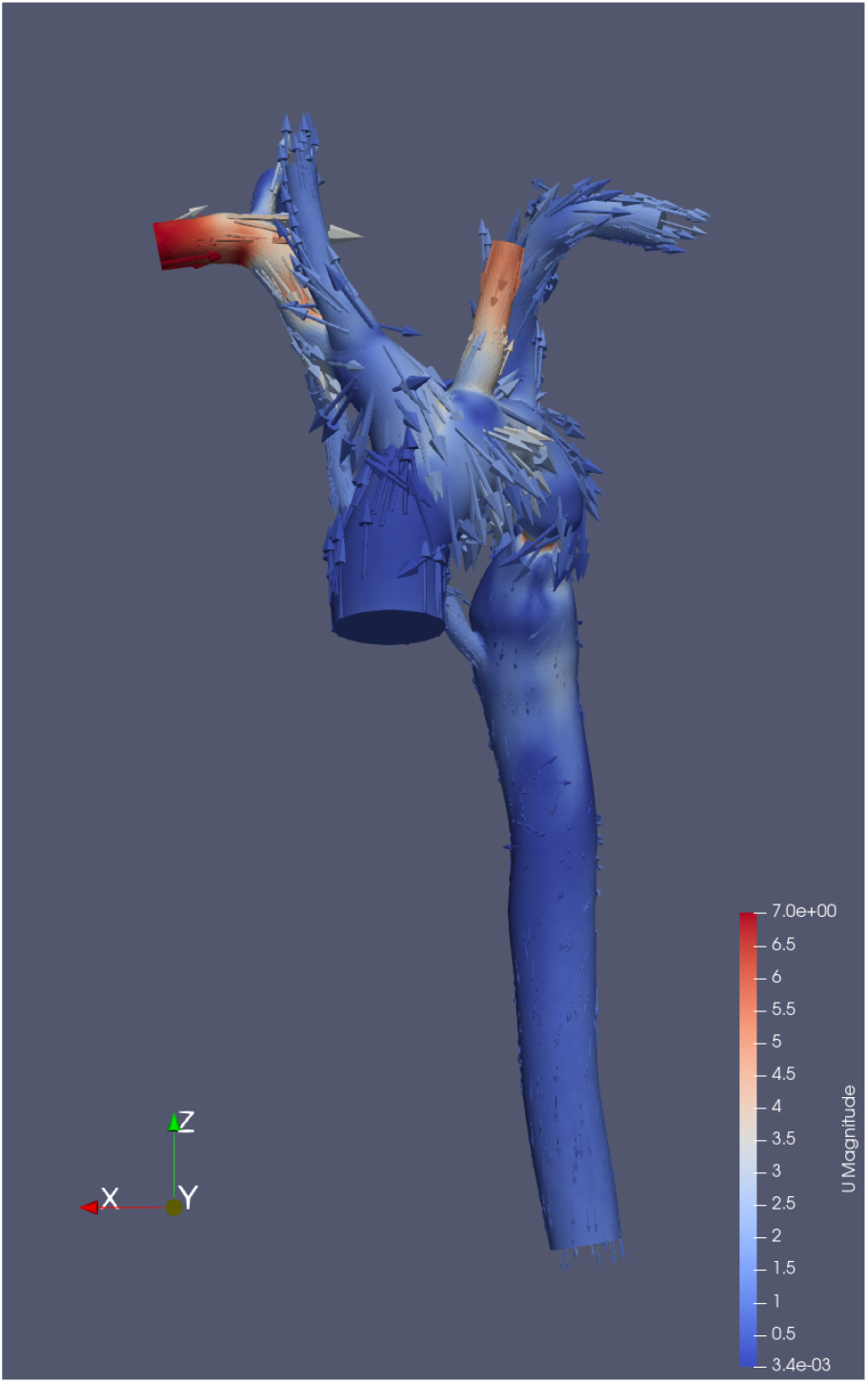
Glyph of velocity at 0.7 seconds. There is clearly significant inflow in the Btrunk face, which is very unexpected. These reversals lead to unexpected results, such as significant inflow through Carotid and Btrunk faces after 0.25 seconds as shown in Figure 13.

**Figure 13:**
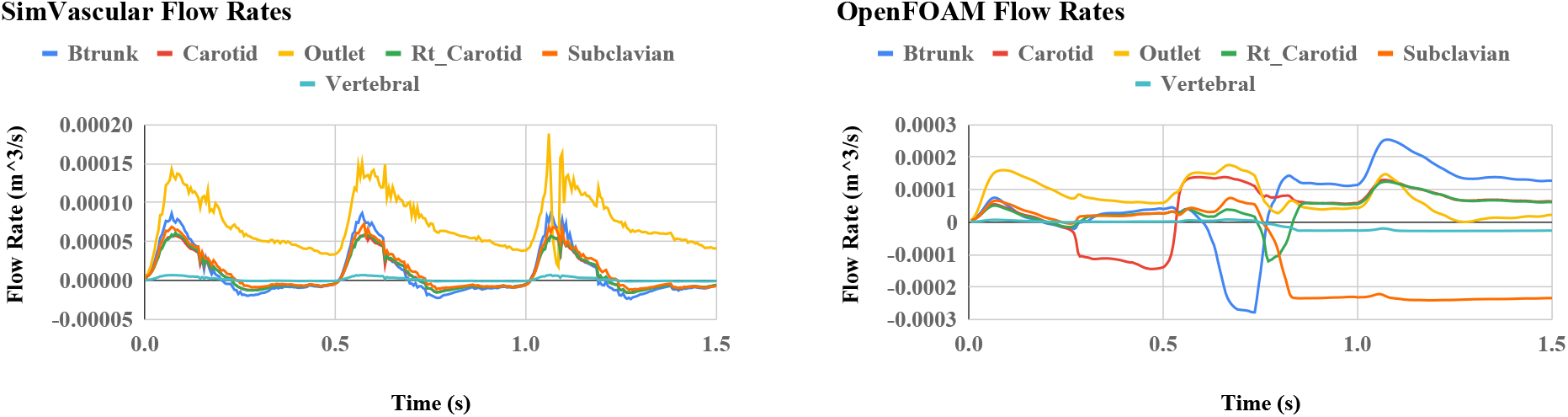
On the left: SimVascular simulated flow rates the outlets. On the right: OpenFOAM simulated flow rates at the outlets. Clearly, there is inflow at unexpected faces at times after 0.25 seconds in the OpenFOAM simulation.
2. The pressure values simulated in SimVascular increase steadily over each period of the inlet flow as shown in Figure 14. This pattern is seen clearly at the inlets and outlets in SimVascular. Since the pressure value from the outlet in SimVascular is used as a boundary condition in OpenFOAM, we can see that OpenFOAM and SimVascular show agreement on the pressure values at the outlets. At the inlet, while the pressure values increase over each period in SimVascular, OpenFOAM maintains the same pressure value levels despite the increase at the outlets. It is clear such periodic increase in pressure is incorrect since it leads to non-sensible values given enough time, it is not clear why the pressure values simulated in SimVascular show such increasing behavior over each period, since the inlet flow rate is exactly periodic as shown in Figure 3. Further investigation is needed to identify the cause of this artificial increase in SimVascular.

**Figure 14:**
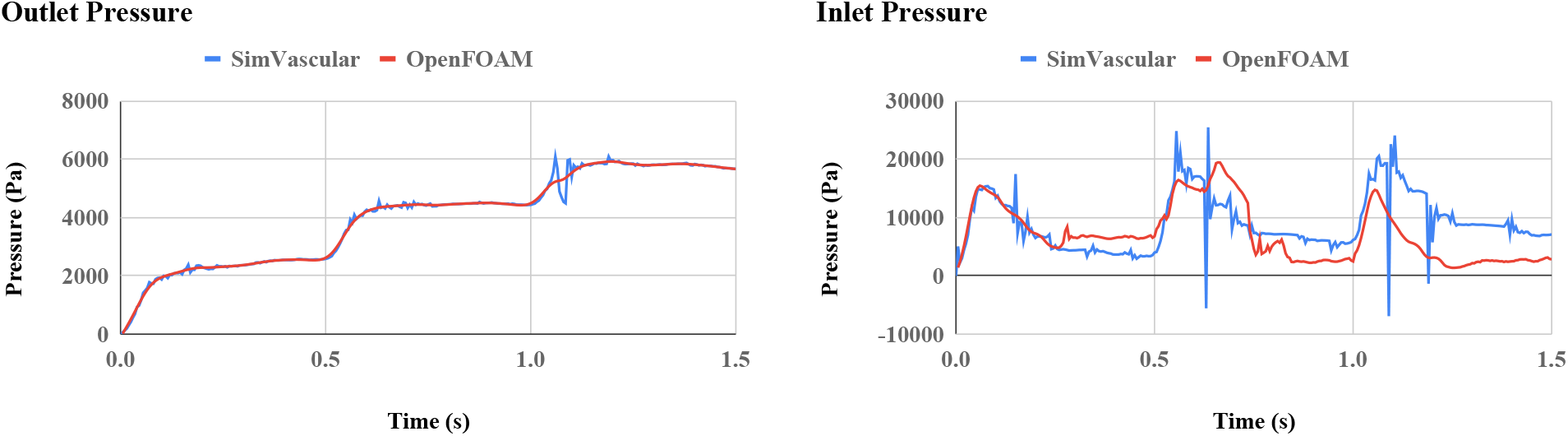
Pressure values simulated in SimVascular over three periods of inlet flow. Both pressures in SimVascular increase every period. The OpenFOAM outlet pressure also increases every period, as prescribed by the boundary condition. However, the calculated inlet flow in OpenFOAM does not increase like that of SimVascular does.
3. The only field that shows significant difference between SimVascular and OpenFOAM during the first 0.25 seconds is WSS. The most plausible cause of this discrepancy is the difference in fluid viscosity model used in the two solvers. It would be useful to implement identical Newtonian descriptions of shear stress formulation and reexamine WSS results from SimVascular and OpenFOAM to verify this hypothesis.
4. The unphysical periodic increase in outlet pressure is applied in OpenFOAM in order to maintain a similar setup with the SimVascular simulation. It would be interesting to also check OpenFOAM results without the artificial increase in outlet pressure.
5. In order to run OpenFOAM without numerical instability, we have to apply a speed limiter to dampen and artificially reduce kinetic energy in the OpenFOAM simulation. The origin of the numerical instability without the speed limiter is not clear in Open-FOAM.
6. Given the velocity, viscosity, and length scale used in the simulation, it would be appropriate to enable turbulence modelling of the blood flow. In this study, both solvers are laminar only due to the lack of support of modelling turbulence in SimVascular. It would be interesting to turn on turbulence in OpenFOAM and compare the results with SimVascular.

We are encouraged by the agreement shown in the results modelled by SimVascular and Open-FOAM. Such agreement gives us confidence to use these tools in haemodynamics modelling. At the same time, there are also many unanswered questions revealed by this preliminary study that could be better answered in future work.

## Data Availability

Data will be made available upon request.

